# Clinical exome sequencing for stroke evaluation uncovers a high frequency of Mendelian disorders: a retrospective analysis

**DOI:** 10.1101/2022.06.10.22276114

**Authors:** Runjun D. Kumar, Linyan Meng, Pengfei Liu, Christina Y. Miyake, Kim C. Worley, Weimin Bi, Seema R. Lalani

**Author notes:** Corresponding Author Seema R. Lalani, One Baylor Plaza, R806, BCM225, Houston, TX 77030.

## Abstract

**Background:** Stroke causes significant disability and is a common cause of death worldwide. Previous studies have estimated that 1-5% of stroke is attributable to monogenic etiologies. We set out to assess the utility of clinical exome sequencing (ES) in the evaluation of stroke.

**Methods:** We retrospectively analyzed 124 individuals who received ES at the Baylor Genetics reference lab between 2012 and 2021 who had stroke as a major part of their reported phenotype.

**Results:** Ages ranged from 10 days to 69 years. 8.9% of the cohort received a diagnosis, including 25% of infants less than 1 year old; an additional 10.5% of the cohort received a probable diagnosis. We identified several syndromes that predispose to stroke such as *COL4A1*-related brain small vessel disease, *CBS*-related homocystinuria, *POLG*-related disorders, *TTC19*-related mitochondrial disease, and *RNASEH2A* associated Aicardi-Goutieres syndrome. We also observed pathogenic variants in *NSD1, PKHD1, HRAS* and *ATP13A2*, which are genes rarely associated with stroke.

**Conclusions:** Although stroke is a complex phenotype with varying pathologies and risk factors, these results show that use of exome sequencing can be highly relevant in stroke, especially for those presenting <1 year of age.

## Background

Stroke is a leading cause of death and long-term disability worldwide. The risk of stroke for children and young adults is lower than for older adults, but is still 1/4,000[1]. Especially for children, “stroke” may include ischemic, hemorrhagic, or metabolic pathology. Not surprisingly, the molecular basis of stroke remains partially understood. Pediatric stroke sometimes occurs as part of an inherited genetic syndrome[2], and can follow Mendelian inheritance patterns[3]. Monogenic causes of stroke include CADASIL, homocystinuria, Fabry disease, *TREX1, COL4A1/COL4A2*-related syndromes, and others[4, 5]. One study of 38 patients with pediatric and perinatal stroke diagnosed 10% of cases using a panel of 15 genes focused on Mendelian causes of stroke[6]. Others have similarly shown the importance of gene-panel testing in evaluation of stroke[7].

To our knowledge, exome sequencing (ES) has not been systematically utilized for evaluation for monogenic causes of stroke[8]. We hypothesized that ES would have high diagnostic rates for infants, children and young adults, with stroke as a presenting complaint. Here we describe the results of ES in 124 individuals with personal histories of stroke referred to our clinical testing facility.

## Materials and Methods

### Clinical Samples

We retrospectively identified 124 patients from over 8,000 clinical samples exome sequenced by Baylor Genetics (Houston, TX) between June 2012 and May 2021. Inclusion criteria included personal histories of stroke or cerebral infarct based on the provided clinical phenotype. Patients who only had family histories of stroke were excluded, as were patients diagnosed by methods besides exome sequencing (e.g. concurrent microarray or mitochondrial DNA sequencing).

There was a single prenatal exome which was retained (Patient ID 87642) with maternal demographic data. Patients provided informed consent for genetic testing, as well as receiving secondary findings or carrier status as recommended by the American College of Medical Genetics and Genomics (ACMG)[9]. The aggregate clinical data were collected and analyzed with the approval of the Baylor College of Medicine Institutional Review Board (protocol H-41191). Patient identification numbers were randomized.

### Exome Sequencing and Analysis

The 124 patients underwent one of proband-only ES (available since December 2011), trio ES (available since October 2014), or critical-trio ES (an expedited test available since April 2015), all of which were performed by Baylor Genetics as clinical assays. Trio ES involves sequencing the proband and both parents, allowing variants to be described as inherited or *de novo*, and in *cis* or *trans*. Critical-trio ES is analytically identical to trio ES, but follows an expedited schedule and is most often used for critically ill patients. A few patients included in this study had Total Blueprint panels, which encompass roughly 5,000 genes associated with Mendelian disease. All samples were concurrently analyzed by Illumina HumanOmni1-Quad, HumanExome-12v1 or HumanCoreExome-24v1 chromosomal microarrays for quality-control. Exome data were interpreted according to ACMG guidelines and variant interpretation guidelines of Baylor Genetics as previously described[10].

### Clinical Exome Reporting

A case was classified as molecularly diagnosed when (1) a pathogenic or likely pathogenic (P/LP) variant was detected, (2) in a gene associated with a syndrome that substantially matched the information submitted to the laboratory, and (3) the zygosity of the variant matched the gene’s established inheritance pattern. Some cases were designated as “probably diagnosed” if there was a variant of unknown significance (VUS) in a gene that substantially matched the reported phenotype, or if there was P/LP variant in a gene that partially matched the reported phenotype. Patients with P/LP variants in genes that cause a known syndrome which does not match the reported phenotype were considered “undiagnosed” for the purposes of this study.

### Statistics

R x64 4.0.3 with base package was used for statistical analyses. Fisher exact tests were used unless otherwise specified.

## Results

### Demographics, Indication, Test Type

Of 124 patients in the study, 67 were male. Ages ranged from 10 days to 69 years. The median age was 9.0 years, with 24 patients younger than 1 year and 74 patients below 12 years. Sixteen patients had metabolic stroke, 9 had prenatal stroke, and 14 had hemorrhagic stroke or stroke associated with vessel dissection. Ninety-three patients had non-neurologic symptoms or findings (Table S1). Eighty-nine patients had proband-only ES, 19 trio ES and 15 critical-trio ES (Table 1). One patient had a total blueprint assay.

**Table 1.** Diagnosis status by test type and patient age. Diagnosed cases had a pathogenic or likely pathogenic (P/LP) variant identified in a gene that causes a syndrome that matched the reported patient phenotype. Probably diagnosed cases had P/LP variants in a gene causing a syndrome that partially matches the observed phenotype, or a variant of unknown significance in a gene causing a syndrome that fully match the observed phenotype.

| Age: | Diagnosis: | Critical-trio ES | Trio ES | Proband ES | TotalBlueprint | Totals |
| --- | --- | --- | --- | --- | --- | --- |
| **<3m** | Undiagnosed | 3 | 2 | 5 | 0 | **10** |
|  | Probably diagnosed | 0 | 0 | 0 | 0 | **0** |
|  | Diagnosed | 0 | 1 | 0 | 0 | **1** |
| **3m-1y** | Undiagnosed | 3 | 3 | 1 | 0 | **7** |
|  | Probably diagnosed | 0 | 0 | 1 | 0 | **1** |
|  | Diagnosed | 2 | 0 | 3 | 0 | **5** |
| **1-5y** | Undiagnosed | 0 | 3 | 14 | 0 | **17** |
|  | Probably diagnosed | 0 | 1 | 1 | 0 | **2** |
|  | Diagnosed | 0 | 0 | 0 | 0 | **0** |
| **5-12y** | Undiagnosed | 0 | 5 | 18 | 1 | **24** |
|  | Probably diagnosed | 1 | 0 | 4 | 0 | **5** |
|  | Diagnosed | 2 | 0 | 0 | 0 | **2** |
| **12-18y** | Undiagnosed | 1 | 2 | 11 | 0 | **14** |
|  | Probably diagnosed | 0 | 0 | 1 | 0 | **1** |
|  | Diagnosed | 1 | 0 | 0 | 0 | **1** |
| **18-40y** | Undiagnosed | 2 | 2 | 10 | 0 | **14** |
|  | Probably diagnosed | 0 | 0 | 2 | 0 | **2** |
|  | Diagnosed | 0 | 0 | 2 | 0 | **2** |
| **>40y** | Undiagnosed | 0 | 0 | 14 | 0 | **14** |
|  | Probably diagnosed | 0 | 0 | 2 | 0 | **2** |
|  | Diagnosed | 0 | 0 | 0 | 0 | **0** |
| **Totals** |  | **15** | **19** | **89** | **1** | **124** |

### Diagnostic Rates

Eleven (8.9%) patients were fully diagnosed; though an additional 13 cases were probably diagnosed, resulting in a 19.4% diagnostic rate when included (Table 1, Table S2). Diagnostic rates varied by patient age. For example, the average age of fully diagnosed cases was 6.7 years, versus an average age of 15.4 for the undiagnosed cases (t-test p-value=0.01, n=11, 100).

Diagnostic rate was highest in infants: 6/24 for patients under 1 year-old versus 5/100 for all other patients (p=0.007, OR=6.2 [95% CI 1.4-28.7]).

The diagnostic rate for critical-trio ES was also higher than proband-only ES at 5/15 versus 5/89 cases fully diagnosed, respectively (p=0.005, OR=8.1 [95% CI 1.6-42.6]). However, critical-trio ES also trended towards outperforming standard trio ES (5/15 versus 1/19, p=0.066, OR=8.4 [95% CI 0.8-447.4]). Since these two tests are analytically identical, we assessed for possible confounding factors.

The average age for patients receiving trio studies was younger than patients receiving proband-only studies (7.1 versus 17.9 years, t-test p<0.0001, n=34, 89). Although trio studies (critical and non-critical) make up 27% of the dataset (34/124), they make up 58% of the tests sent in infants less than 1 year-old (14/24, p=0.0007, OR=5.2, [95% CI 1.8-15.1]). Test type and age are sufficiently confounded that we could not ascribe a separate diagnostic rate to trio testing versus proband-only after correcting for age.

### Diagnoses

A P/LP variant that substantially matched the reported phenotype was identified in 11/124 cases, (Table 2). Six solved cases were under 1-year-old, with diagnoses that included Aicardi-Goutieres and *POLG*-related syndromes. Additionally, in six solved cases the gene was previously associated with stroke, based on reviews of OMIM, GeneReviews and literature searches. Among that group are patients 31162 with *CBS*-related homocystinuria (MIM# 236200), patient 84711 with *TTC19*-related mitochondrial complex III deficiency (MIM #615157), and patient 87229 with *POLG*-related mitochondrial disease.

**Table 2.**
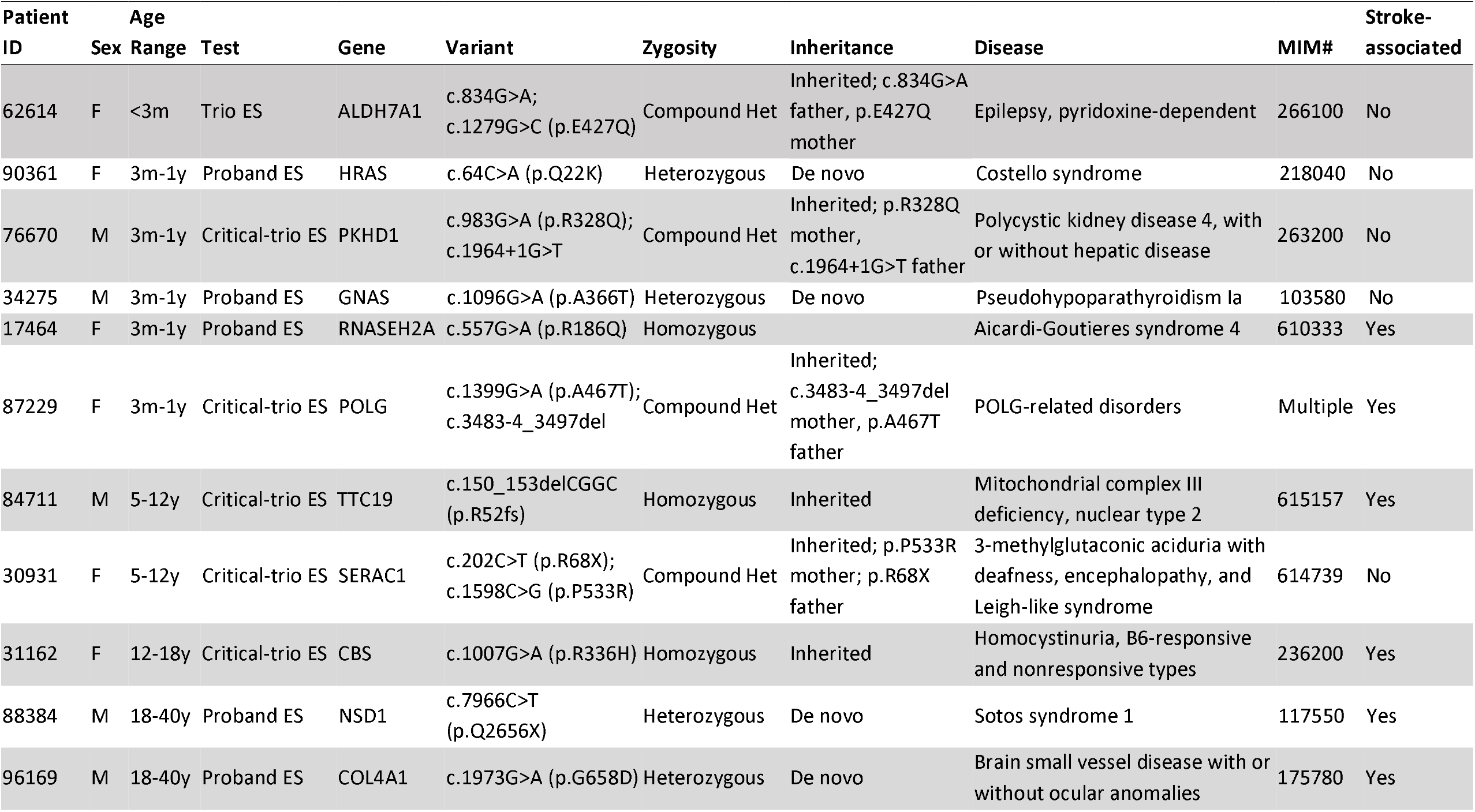
The 11 fully diagnosed cases. The pathogenic or likely pathogenic variant and the affected gene are shown, as well as additional data that were clinically reported.

### Recurrent genes observed in the cohort

No genes recurred among the 11 diagnosed cases. However, an additional 13 cases were designated as “probably diagnosed” (Table S2). Including these additional cases, the only recurrent genes were *COL4A1* and *ATP13A2*.

*COL4A1* causes several autosomal dominant syndromes that have been associated with stroke[6]. Patient 96169 was an adult male with developmental delay, hypotonia, seizures, dysmorphic facies, cataracts and brainstem strokes. Brain MRI was concerning for agenesis of the corpus callosum, porencephalic cysts, and cerebellar hypoplasia. Through proband ES, he was diagnosed with brain small vessel disease with or without ocular anomalies (BSVD1, MIM# 175780, Table 2). Two additional patients were considered probably diagnosed for this condition (Table S2). Patient 87642 was a fetus with intrauterine growth restriction, microcephaly, microvascular proliferation of the retina, congenital heart disease, and brain MRI consistent with cerebral hemorrhages and infarcts. Proband ES by amniocentesis showed a c.2879G>T (p.G960V) variant in *COL4A1* which was called likely pathogenic for BSVD1. Patient 71712 was a 2-to-5 year-old girl with developmental delay, intellectual disability, seizures, dysmorphic features, microcephaly, congenital cataracts and stroke-like episodes. A novel *de novo* heterozygous c.3555A>G (p.K1185K) VUS was identified in *COL4A1* and predicted to affect splicing.

Two patients were considered probably diagnosed with variants in *ATP13A2*. This gene is associated with Kufor-Rakeb syndrome and spastic paraplegia 78 (MIM# 606693, 617225), autosomal recessive disorders associated with juvenile and adult-onset Parkinsonism, respectively, although neither is associated with stroke. Patient 51411 was an adult male with seizures, apraxia, memory problems, confusion and progressive cognitive decline and history of stroke; proband-only testing revealed a heterozygous c.348-9_351del pathogenic splice-site variant and c.943G>A (p.G315R) VUS of *ATP13A2*. The phase of these variants could not be determined. Patient 20796 was a male infant with seizures, microcephaly, failure-to-thrive, and cataracts. Brain MRI showed evidence of cerebral atrophy, periventricular leukomalacia, and stroke. Proband ES showed homozygous c.3056delG (p.G1019fs) variant in *ATP13A2*.

### Syndromes not previously associated with Stroke

In our study, 5/11 fully diagnosed cases involve disorders that, to our knowledge, have not been previously associated with stroke (Table 2). We will highlight two of these cases.

Patient 90361 was a female infant with intrauterine stroke, delayed motor milestones, dyskinetic movements, dilated aorta and acquired obstructive hypertrophic cardiomyopathy. Proband ES revealed a *de novo* heterozygous c.64C>A (p.Q22K) pathogenic variant in *HRAS* which is associated with autosomal dominant Costello syndrome (MIM# 218040). Costello syndrome can cause growth failure, intellectual disability, and dysmorphic facial features in addition to other skin and musculoskeletal anomalies. There is no known association between Costello syndrome and stroke, though related disorders Noonan and Neurofibromatosis 1, which also affect the Ras signaling pathway, have been reported with stroke[11, 12].

Patient 76670 was a male infant with premature birth, oligohydramnios, anisocoria, seizures, polycystic renal dysplasia, and recurrent vascular ischemic strokes. Critical-trio ES revealed heterozygous pathogenic c.983G>A (p.R328Q) and c.1964+1G>T changes in *PKHD1* which were inherited in *trans*. Defects of *PKHD1* are a cause of autosomal recessive polycystic kidney disease (ARPKD, MIM# 263200). ARPKD typically presents in the neonatal period with oligohydramnios, massively enlarged kidneys, with perinatal death in ∼30% of affected newborns. Although autosomal dominant polycystic kidney disease can present with hemorrhagic stroke in young adulthood, the recessive forms have been associated with nonbleeding aneurysms[13], or rarely with optic nerve ischemia[14]. Stroke due to sequelae of hypertension cannot be excluded.

## Discussion

This study represents one reference laboratory’s experience using clinical ES for patients with stroke. Our study indicates that patients with stroke of all ages, and especially patients with perinatal or pediatric stroke are likely to benefit from ES. We diagnosed nearly 9% of cases, and 25% of infants under the age of 1; an additional 10% of cases received probable diagnoses. In our cohort, *COL4A1*-related brain small vessel disease was the most frequent diagnosis. Mitochondrial diseases due to biallelic variants in *POLG* and *TTC19* were found in two children in our study, presenting with ischemic stroke and multiple stroke-like episodes respectively.

Variants in *TTC19* cause complex III deficiency and have recently been shown to cause recurrent stroke-like episodes with basal ganglia changes[15]. Homocystinuria due to *CBS* variant and Aicardi-Goutières syndrome resulting from *RNASEH2A* variants were also identified in our study. Interestingly, we identified a subject with Sotos syndrome due to a pathogenic *NSD1* variant, who had a stroke at age 18 years. Sotos syndrome is an overgrowth syndrome which is not classically associated with stroke, however it has recently been associated with arterial thrombosis, hemorrhagic stroke, and subdural hematoma in rare patients[16]. Our data indicate that *NSD1* evaluation should be included in the genetic work up for stroke.

Importantly, ES also identified pathogenic variants in genes not currently known to increase the risk of stroke. Beyond *PKHD1* and *HRAS* discussed earlier, our study found a subject with a neurodegenerative disorder affecting mitochondrial function, *SERAC1*-related MEGDHEL syndrome (3-methylglutaconic aciduria with deafness-dystonia, hepatopathy, encephalopathy, and Leigh-like syndrome), which is not recognized to be associated with stroke at present.

Further studies are needed to understand the relevance of *SERAC1* with stroke, if any.

There are some important limitations to our study. Stroke is a challenging phenotype for genetic analysis. Foremost, “stroke” may represent a number of pathologies, such as hemorrhage or ischemia, and our dataset had limited clinical details to separate these entities. Especially in the pediatric population, the presence of infection, vasculopathy, congenital heart disease, coagulopathy and other risk factors may influence stroke risk irrespective of the genetic determinants. However, clinicians often must order testing without access to these data. The perinatal period is an excellent example, where the original insult can occur prior to delivery with limited characterization. Our analysis indicates that ES is particularly well suited to assessing stroke in the first year of life.

Given recent guidance encouraging the use of exome and genome sequencing, it is highly likely that the number of patients undergoing these genetic assays will increase rapidly[17]. Ongoing analysis of the generated data will be essential to assessing stroke risk for patients with previously described genetic disorders, and eventually to help identify new heritable stroke syndromes.

## Conclusions

In this study we retrospectively analyzed 124 patients with personal history of stroke referred for exome sequencing. Nearly one fifth of patients received a complete or probable diagnosis, and one quarter of patients less than one year old were fully diagnosed. Several individuals have Mendelian disorders that are not typically associated with stroke, including Sotos syndrome, autosomal recessive polycystic kidney disease, and Costello syndrome. This analysis indicates that patients with stroke, especially at younger ages, are likely to benefit from broad genetic testing.

## Data Availability

All data described in this study are provided within the article and supplementary materials. Additional de-identified clinical data is available upon request to the corresponding author.

## Declarations

### Ethics Approval

This study adheres to the principles set out in the Declaration of Helsinki. The study was approved by the Baylor College of Medicine Institutional Review Board (IRB protocol H-41191).

### Author Contribution

Conceptualization: R.D.K., S.R.L.; Data curation: R.D.K., S.R.L., L.M., P.L., K.C.W.; Formal analysis: R.D.K.; Writing – original draft: R.D.K., S.R.L.; Writing – review & editing: R.D.K., S.R.L., C.Y.M., L.M., P.L., K.C.W.

